# Phylogeny-based estimates of epidemic population substructure outperform time series approaches

**DOI:** 10.1101/2025.01.28.25321284

**Authors:** Julie A. Spencer, Emma E. Goldberg, Prescott C. Alexander, Sara Y. Del Valle

## Abstract

Improving infectious disease models plays a crucial role in mitigating global health impacts. Population-level models have been essential for understanding and anticipating impacts of infectious diseases, but their accuracy has often been limited due to changing environments, rapid pathogen evolution, and insufficient access to detailed transmission data. In particular, due to the lack of granular transmission data, many epidemiological models assume homogeneous disease transmission rather than acknowledge differences between groups within the host population. To address this gap, we conducted a simulation study to assess the potential for inferring group-structured viral transmission dynamics from phylogenetic data and to demonstrate how age structure can be incorporated in case projections. Using a synthetic dataset of 800 age-structured viral phylogenies and their associated case time series, we estimated transmission rates within and between age groups. Our results show that the estimates derived from phylogenies are more accurate than those from case counts alone, although both approaches enabled recovery of age-specific transmission differences. These results suggest that viral phylogenies and case-count time series each provide valuable data that can improve our understanding of population-level disease dynamics, ultimately contributing to more accurate and effective models for informing public policy and reducing disease burden.

## 1 Introduction

Accurate and timely models are essential tools for anticipating the emergence and spread of infectious diseases and protecting global communities from their impacts [1, 2]. Epidemiologists have shown that incorporating heterogeneous population substructure into disease models improves their accuracy for mitigation and forecasting, as transmission dynamics and susceptibility often vary by group [3, 4, 5, 6]. Here, we focus on age structure as an example of this form of heterogeneity.

Age-specific transmission matrices are critical for models of diseases like COVID-19, where age is a major risk factor for severe outcomes. These models can help decision-makers evaluate interventions such as school closures, business restrictions, and targeted vaccination strategies [7, 8]. However, data for informing age-structured transmission are often scarce due to limited knowledge of the timing and direction of individual infection events, leading many epidemiological models to assume homogeneous transmission [9].

Age-structured models are not only essential for diseases like COVID-19, but they have also been instrumental in understanding childhood infectious diseases, particularly in the context of vaccination prog. Models incorporating age-specific contact patterns have played a critical role in optimizing vaccination strategies for diseases like measles, mumps, and rubella, where childhood vaccinations significantly impact population immunity and disease transmission dynamics [10, 11]. For instance, age-structured models have informed the design of childhood immunization schedules and have helped in understanding the indirect effects of vaccines on different age cohorts through population immunity [12, 13]. These models highlight the long-term effects of vaccination programs, including waning immunity and the need for booster shots, further emphasizing the importance of accurate age-structured data [14, 15].

During the past 20 years, researchers have made substantial progress in characterizing the age heterogeneity of host populations and integrating that information into epidemiological models [16]. A persistent challenge, however, has been the availability of high-quality data to accurately parameterize these models. Age-structured SIR-type models, network-based models, agent-based models, and various statistical methods rely on case reports, demographic data, social contact surveys, clinical records, serological data, and mobility data [17, 18, 9, 19, 5, 20, 21]. Each of these datatypes has its own limitations such as under-reporting, recall bias, or a lack of granularity in capturing individual-level transmission events.

Due to advances in sequencing and computing technologies, pathogen genomic sequences in public repositories present one rapidly increasing data source [22, 23]. Specifically, phylogenetic trees inferred from pathogen genomes can provide critical information about timing and direction of disease transmission, making them amenable to new methods for inferring age structure [24, 25, 26, 27]. However, challenges such as low sampling rate (a small proportion of sequenced samples to total actual cases) continue to limit their broad application [28].

In this study, we demonstrate how age-structured transmission rates can be estimated from either viral phylogenies or case count time series alone. To achieve this, we conduct a simulation study with synthetic datasets of viral phylogenies and corresponding case time series generated under four scenarios of age-dependent transmission. By fitting models of exponential epidemic growth to each dataset, we show that both data types contain information about the age-stratified transmission process, but that the estimates from the phylogenies are more accurate and less sensitive to stochastic effects in data generation (see Figure 2). Although age structure can provide important information about an outbreak, we further show, using time series-based parameter estimates, that case counts can be projected equally well without this information.

This proof-of-concept study reveals the potential for viral phylogenies and case counts to jointly advance our understanding of heterogeneous transmission dynamics. Additionally, our approach has broad applicability to diseases where age plays a crucial role, including not only COVID-19 but also influenza, respiratory syncytial virus (RSV), and other respiratory illnesses.

## 2 Methods

### 2.1 Epidemiological process

Our approach involves simulating a simple epidemiological process in which transmission rates differ for different groups. For concreteness, we model two age classes—“adult” and “child”—and generate datasets representing viral phylogenies and time series of cases. We then assess the accuracy of inferring age-structured transmission rates from each data type.

Because our intent is primarily to compare the extent to which age-structured transmission is encoded in the two data types, we make several simplifying assumptions that apply to all the simulated data. These include modeling the epidemic’s exponential growth phase as a basic dynamical system, which enables clearer isolation of age-structured transmission patterns in our analysis (see Figure S2). Using these assumptions, we arrived at the following system of differential equations:

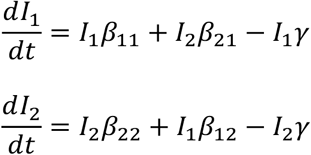

Our model focuses on the infected classes, *I*_*i*_, and assumes that the susceptible population is unlimited, unaffected by depletion due to the ongoing outbreak. The parameters *β*_*ij*_ represent the transmission rates from individuals in class *i* to those in class *j*, while *γ* denotes the rate at which individuals become non-infectious, incorporating possible outcomes such as recovery, isolation, or death.

We further assume that all cases become non-infectious at the time they are sampled. Using the **sim.bdtypes.stt.taxa()** function from the R package TreeSim [26], we simulate transmission trees under the age-structured exponential growth process shown by Eq. 1. For each scenario, we simulate 200 transmission trees, each consisting of the first 200 cases in an outbreak. The proportion of cases in each age group as well as the duration of the outbreak vary stochastically and are functions of the initial conditions and transmission rates in each scenario. To explore the impact of age-structured transmission, we consider four distinct scenarios for values of the transmission parameters *β*_*ij*_:

1. **Homogeneous transmission**: Transmission rates are independent of age group.
2. **Increased within-group transmission**: Transmission rates are higher within age groups than between groups.
3. **Asymmetric transmission**: One age group has higher transmission rates to both groups than does the other group.
4. **Extra asymmetric transmission**: One age group has higher transmission within-group. The other age group, as well as between-group rates, remain lower.

These scenarios span a variety of patterns for how age-structured transmission can influence the resulting epidemiological and phylogenetic data. Parameter values for each of these scenarios are given in Table 1.

**Table 1:**
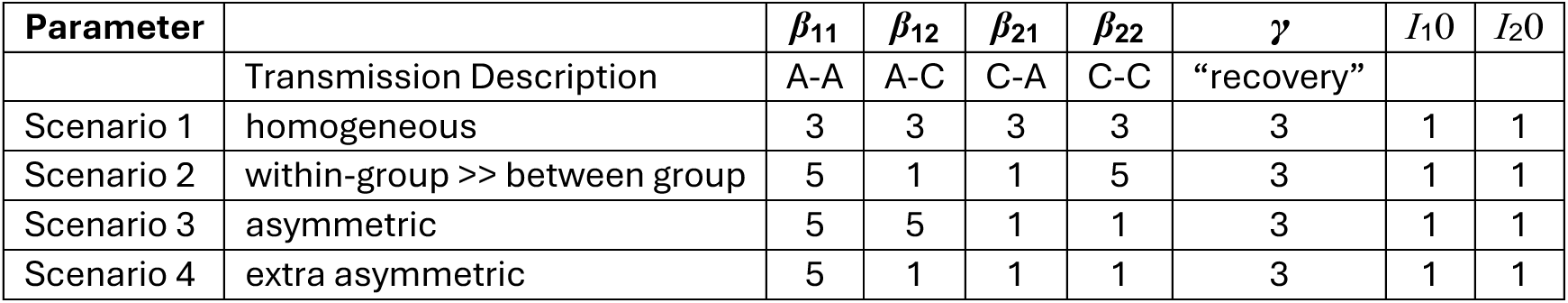
Parameter values used to simulate data under four scenarios of age-structured transmission. As defined in Eq. 1, the *β* parameters represent transmission rates for age groups designated as A (Adult) and C (Child), and *γ* represents the rate of becoming-non-infectious. *I*_1_ represents infected adults and *I*_2_ represents infected children. Initial conditions for all scenarios are *I*_1_0 = *I*_2_0 = 1.

### 2.2 Phylogenetic data and parameter estimation

To convert each simulated transmission tree into a viral phylogeny, we make two simplifying assumptions: (1) the within-host viral diversity is sufficiently low to disregard any pre-transmission interval [29]; and (2) the viral mutation rate is high enough to generate sufficient mutations to resolve the branches of the phylogeny. That is, we assume that the viral phylogeny exactly mirrors the transmission tree, although lacking the information of who infected whom (Fig. 1). Note that **sim.bdtypes.stt.taxa()** does not return who infected whom as node states, so we use the simulated transmission trees directly as viral phylogenies.

**Figure 1:**
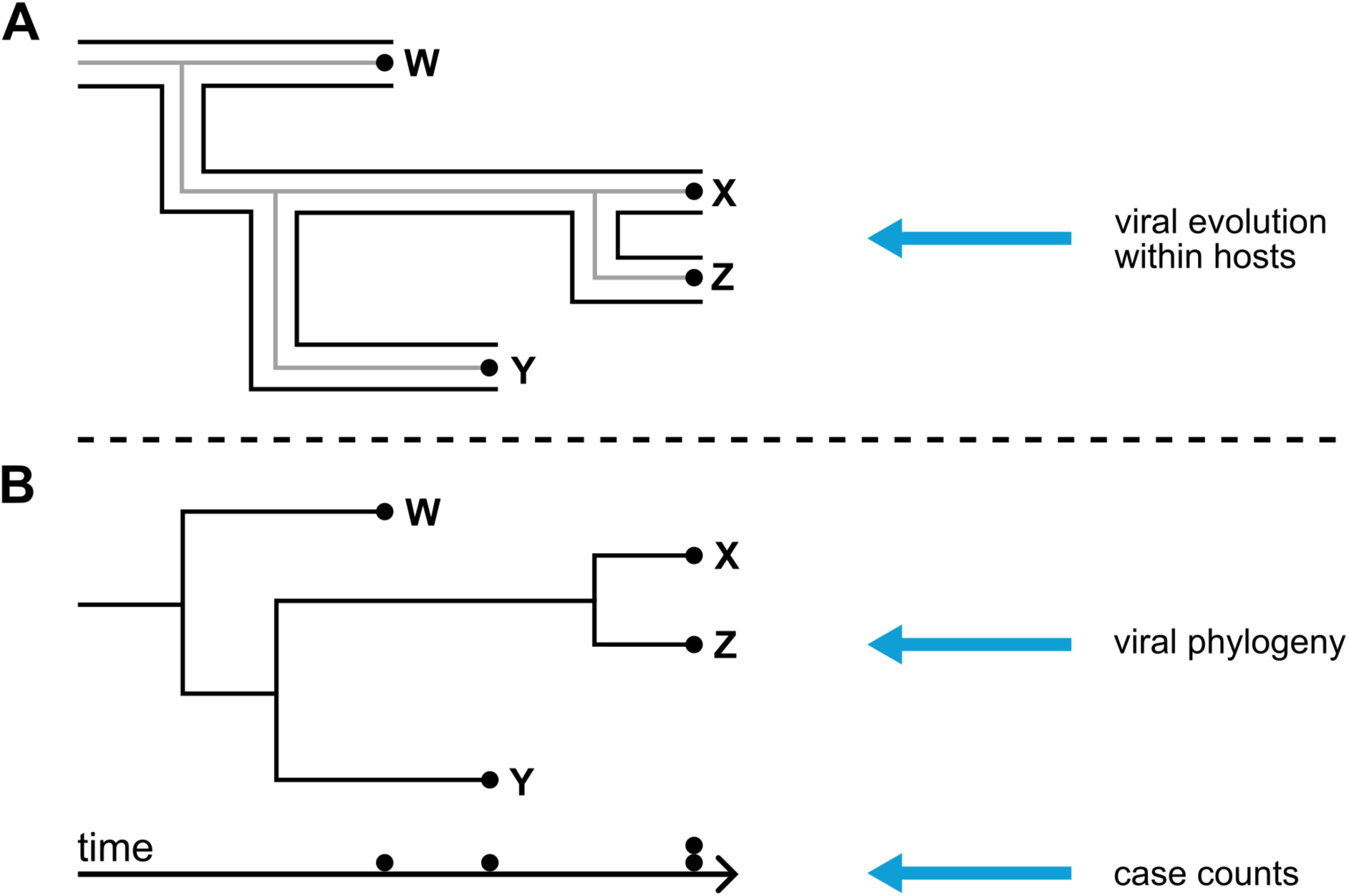
Cartoon of the synthetic data generation process. (A) A transmission tree shows who infected whom, when infections occurred, and when individuals were sampled. This epidemiological process is depicted with the broad outlines for hosts *w*, *x*, *y*, and *z*. Inside these hosts a virus evolves, shown by the internal gray tree, which reflects the simplifying assumption of ignoring within-host viral diversity. (B) When viral genetic sequences are collected, here with one sample per host, a viral phylogeny can be constructed. The phylogenetic tree shows which samples are more closely related, but not directly who infected whom. The case count time series includes only the information about sampling times, not about relatedness (Fig. 2).

**Figure 2:**
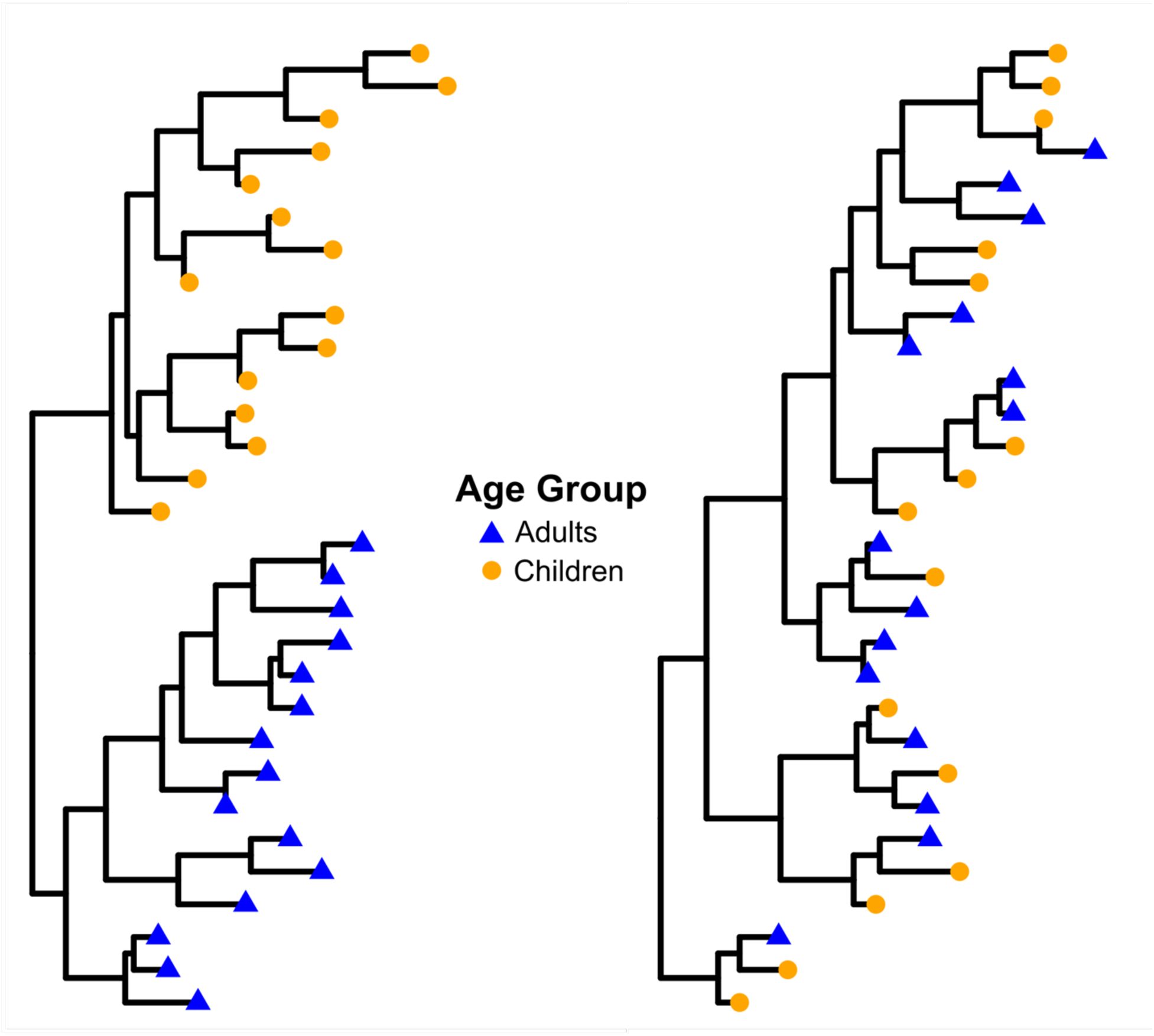
Viral phylogenies contain information about age-structured transmission. For data generated with more within-group transmission (left), more closely related cases tend to be within the same age group. For data generated with more between-group transmission (right), closely related cases are more often from different age groups. Note that for each scenario, age structure in the time series of case counts is not apparent.

For each simulated viral phylogeny, we estimate transmission parameter values by fitting a two-state birth-death process using the **LikTypesSTT()** function in the TreePar package [30], maximizing the likelihood using the **optim()** function in R Statistical Software (v4.4.1) [31]. The birth-death process, modeled as a continuous-time Markov process, captures the key events of transmission (births) and individuals becoming non-infectious (deaths, corresponding to terminating branches). Becoming non-infectious can represent recovery, isolation, death, or sampling. Since the sampling probabilities are set to one, this inference model is equivalent to the data-generating model.

### 2.3 Case time series data and parameter estimation

To convert each simulated transmission tree into a time series of cases, we record the sampling time of each tip, which corresponds to the time that each case was sampled. For computational tractability during model fitting, we bin the cases into time intervals of integer units.

We then estimate transmission parameters for each simulated time series by fitting a two-state ODE model, as described in Eq. 1, to the time series data using the R package **fitode** [32]. Using only the case time series as data preserves the underlying dynamics but removes information on the relatedness of samples that is available for parameter estimation (Fig. 1B).

### 2.4 Effect of age structure on case projections

To show the effect of age structure on case count projections, we generate one additional, larger data set of 1000 cases. The first 300 cases are used for fitting model parameters, and the last 700 cases are used to evaluate model projections.

Using only the stochastically generated training data (the first 300 cases described above), we estimate parameter values under the deterministic age-structured model for the time series, following the methods previously outlined. This fitted, deterministic model is then simply projected forward (numerically simulated using the **predict** function from the **fitode** R package [32]) through the testing period to project case counts.

To compare these against projections that ignore age structure, we additionally fit a non-age-structured exponential growth model, 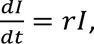 to the same (but unstructured) training data where the initial guess for *r* = 2. This simpler model includes a single parameter, *r*, representing the difference between the transmission rate and the rate of individuals becoming non-infectious. The data given to this model are the total cases per day, the sum of adult and child cases. Case count projections for the testing period are generated via numerical simulation using the fitted parameter values from the training period.

The performance of the two models is compared by evaluating how accurately their projections match the observed case counts in the testing data, using Mean Absolute Error (MAE). This comparison evaluates the potential impact of incorporating age-structured transmission dynamics on the accuracy of outbreak forecasting.

## 3 Results

### 3.1 Parameter estimation results

The parameter estimates derived from synthetic phylogenies consistently outperformed those from the time series in terms of accuracy as shown in Figs. 3, S1. In Scenario 1, both methods exhibited relatively homogeneous transmission rates between and among age groups, clearly reflecting the basis on which this scenario was created. Similarly, in Scenario 2, both methods successfully reproduce the generating transmission pattern, i.e., higher transmission within groups. In Scenario 3 as well, both estimation methods capture the higher transmission from adults to adults and from adults to children. Scenario 4, characterized by extra asymmetric transmission, showed that both methods could capture the greater transmission within adults, contrasted with the other three transmission rates.

**Figure 3:**
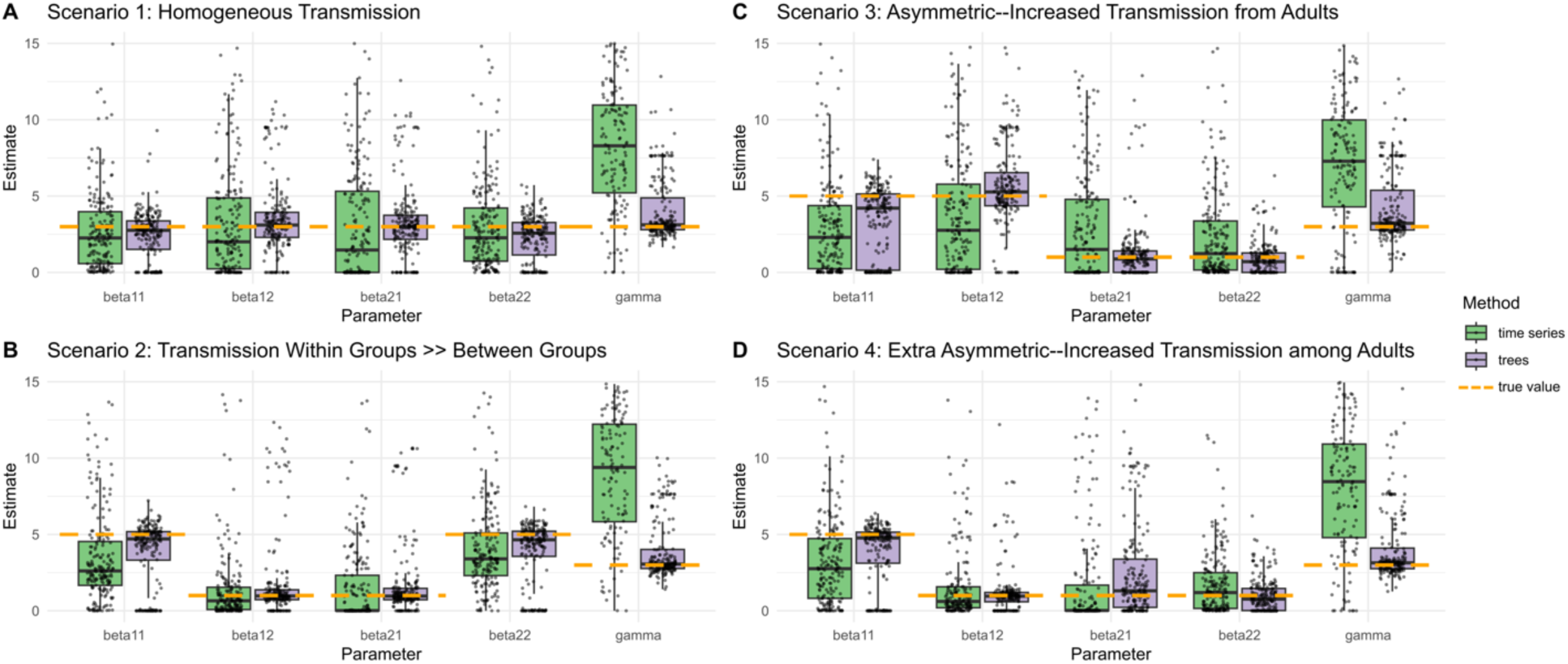
Maximum likelihood parameter estimates from synthetic data, comparing phylogenies (purple) and their corresponding time series (green) across four transmission scenarios. **Panel A** shows Scenario 1, where homogeneous transmission rates are correctly inferred, with estimates similar to the true values used to generate the trees. **Panel B** illustrates Scenario 2, where estimates correctly recover lower between-group transmission, while **Panel C** (Scenario 3) accurately reflects higher transmission rates from adults compared to those from children. **Panel D** (Scenario 4) shows that the pattern of higher transmission rates within adults compared to the other three rates are successfully recovered. The horizontal dashed orange lines indicate the “true” parameter values used to simulate both phylogenies and time series, given in Table 1. The narrower confidence intervals in the phylogenetic estimates highlight the reduced sensitivity to stochastic noise, in contrast to the broader intervals seen in the time series estimates. This figure shows overall better accuracy of phylogenetic data for parameter inference, particularly in scenarios involving structured transmission dynamics.

The spread in the variation of the 200 replicates in each scenario reinforces these results (Fig S1). Estimates derived from time series show consistently larger variation compared to those from phylogenies, reflecting greater uncertainty in parameter recovery from time series data. Please see Supplementary Material available on Cambridge Core website for additional information.

Although the parameter estimates from synthetic viral phylogenies were more accurate in every respect than were the estimates from time series, the time series estimates were still able to recover the broad patterns of transmission heterogeneity across all scenarios (Fig. 3). This suggests that while phylogenetic data provides more precise inference of transmission parameters, time series data can still capture group-structured transmission dynamics at a coarser level when analyzed with an appropriate model.

#### Effect of age structure on case projections

Comparisons of structured and unstructured projections, with both types of projections based on fits to synthetic time series data, yielded Mean Absolute Error (MAE) distributions represented by the boxplots in Figure 4. In each scenario, 200 MAEs are shown for each method. Keeping in view our assumptions and models discussed in the Methods section of this paper, although the distributions are broad, projection accuracy seems to be robust to group structure, at least during the exponential phase of an outbreak. The slightly higher median MAEs for structured model in Scenarios 1-3 may be the effect of fitting deterministic models to stochastically generated data. Please see Supplementary Material available on Cambridge Core website for sample time series projection plots (Figures S3, S4, S5, S6).

**Figure 4:**
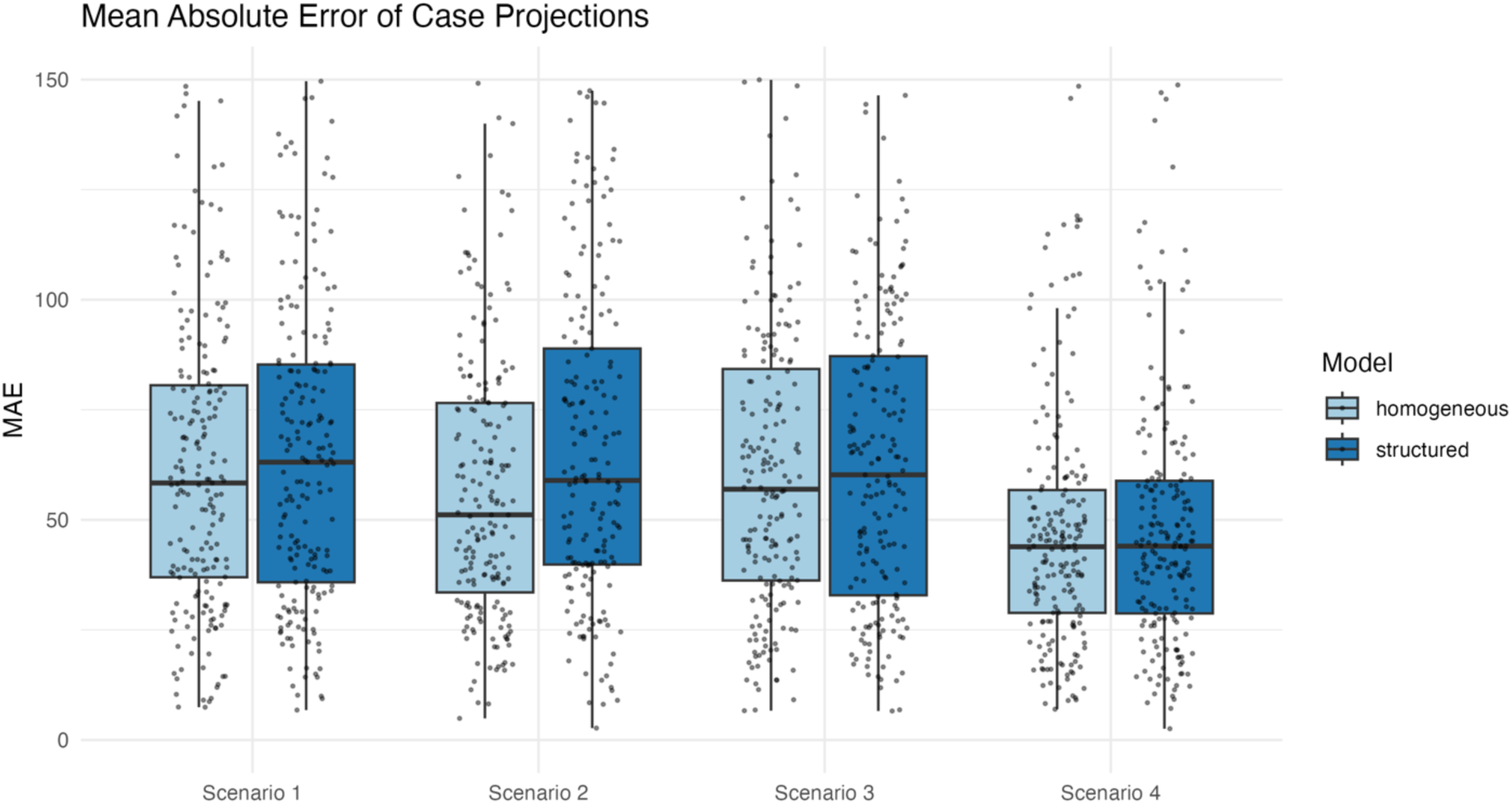
Distribution of the Mean Absolute Error (MAE) for 200 replicates of case projections in the four scenarios studied, using the age-structured model (dark blue, right boxplots) and the unstructured model (light blue, left boxplots). Distributions of MAEs for both the age-structured projections and unstructured projections are broad, likely reflecting the fitting and projection process of deterministic models against stochastically generated data.

## 4 Discussion

In this study, we used synthetic data to evaluate the accuracy of estimating age-structured transmission rates using two different data types: viral phylogenetic trees and case count time series. We carried out these analyses across four scenarios representing age-dependent differences in transmission dynamics. Our results show that parameter estimates derived from phylogenetic trees are more accurate than those from time series, likely because phylogenies encode information about viral genetic relatedness, which offers information about transmission pathways not available from time series alone. At the same time, both approaches enable recovery of the overall transmission heterogeneity embedded when synthesizing the data. Despite the underlying group-structured dynamics, we found that in all scenarios, including those in which transmission was asymmetrical within and between age groups, case projections fitted using an age-structured model have similar accuracy to projections using an unstructured model. Nonetheless, if were to consider individual, stochastically generated data sets that show strong age structure in their parameter estimates, it is possible that age-structured model projections would perform better on these data sets (e.g. see Figure S4).

By using synthetic data with an assumed 100% sampling probability, we avoid challenges associated with incomplete sampling such as information loss and bias. However, in real-world situations, viral sequences are typically available for only a subset of cases, which could limit the utility of phylogenetic approaches. The statistical power of the time series-based approach may be greater than that of phylogenies if case time series are more complete. Moreover, our simulations also assume error-free viral phylogenies, whereas in practice phylogenetic reconstructions are subject to uncertainties. Combining informative, less-sampled phylogenies with less informative but more complete time series could provide a more comprehensive picture of transmission heterogeneity [33, 1, 34, 35, 23].

Our proof-of-concept simulation study is limited to modeling only the exponential phase of an outbreak, which assumes unlimited susceptible individuals and unlimited infection potential, and does not consider the infectious peak, decline, or subsequent oscillations. If additional model dynamics beyond the exponential phase were considered, parameter estimates and comparisons could have different results. We assume homogeneity and constant transmission rates within groups, which are likely not realistic assumptions. We also do not consider selection on viral genotypes for increased transmissibility, regardless of age group. How these and other real-world complexities affect the detection of group-structured transmission remains to be seen.

Our approach can be used for further exploration into how transmission heterogeneity manifests in other population substructures—such as those defined by geography, occupation, or social networks [36]. Additionally, the incorporation of real-world complexities such as population-level interventions (e.g., vaccination or physical distancing) into these models could provide insights into the effectiveness of public health strategies during outbreaks.

The estimates from synthetic trees and from synthetic time series each correctly recovered the age-structured transmission differences present in the data. This suggests that we can capture information regarding heterogeneity from both types of data. Phylogenies as a newly abundant data source show promise in their ability to improve parameter estimates. This knowledge can improve our understanding of population-level disease dynamics, ultimately contributing to more accurate and effective models that can inform public policy and reduce disease burden.

## 5 Acknowledgments

We are grateful to the LANL PreCog team for insightful discussions and to David J. Butts for helpful input on code design.

## 8 Data Availability

Data and workflows for this study are generated from code publicly available in the repository at https://github.com/lanl/precog/tree/main/aim.

## Supplementary Material

**Figure S1.**
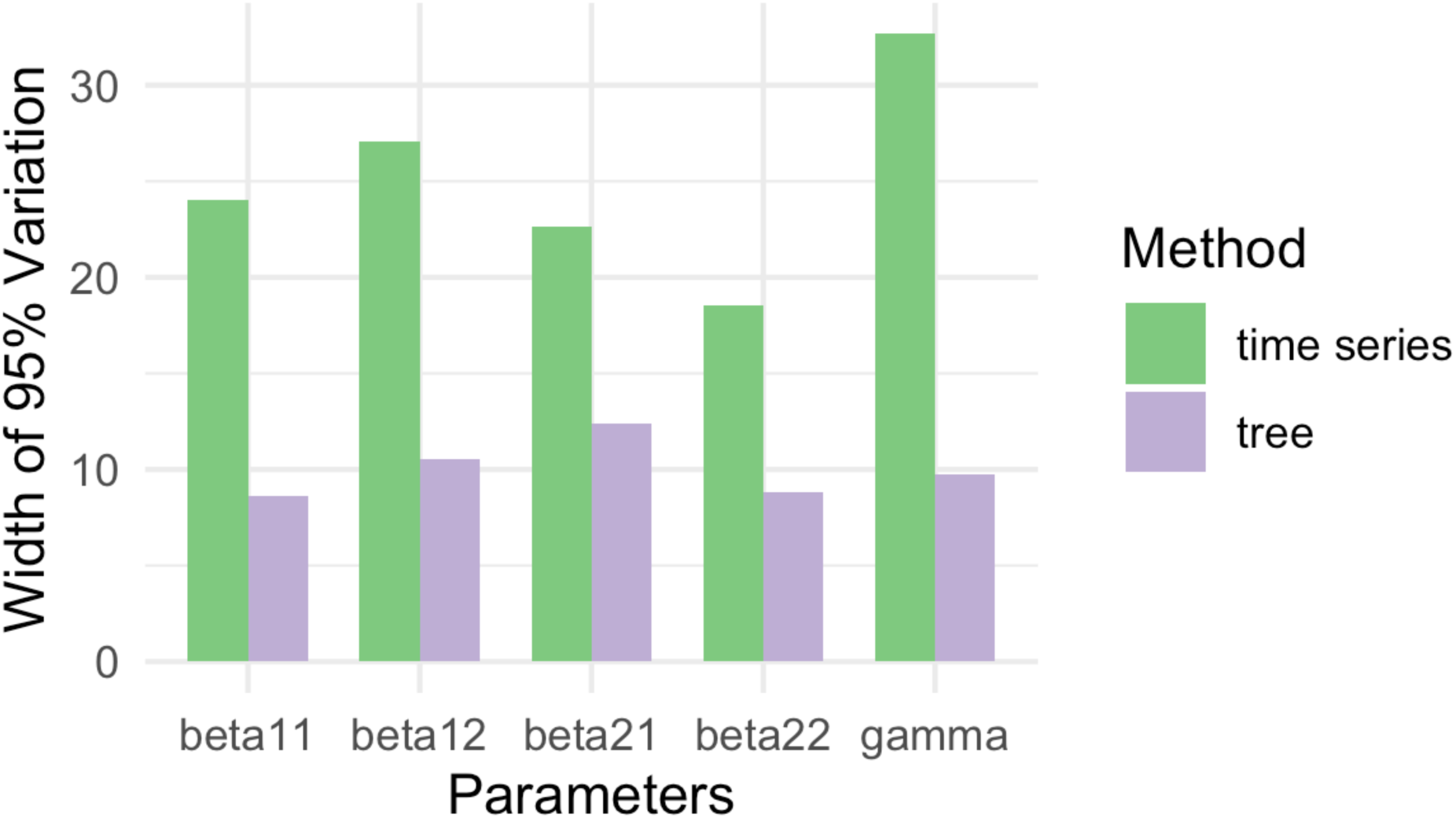
Figure 5: Spread of the variation in 95% of the data from maximum likelihood parameter estimates from 200 synthetic phylogenies (purple) and 200 synthetic time series (green). The parameters are labeled on the x-axis and the widths framing the spread of the estimates (97.5th% and 2.5th% percentiles) is shown on the y-axis. Estimates derived from time series show consistently larger variation compared to those from phylogenies, reflecting greater uncertainty in parameter recovery from time series data.

**Figure S2.**
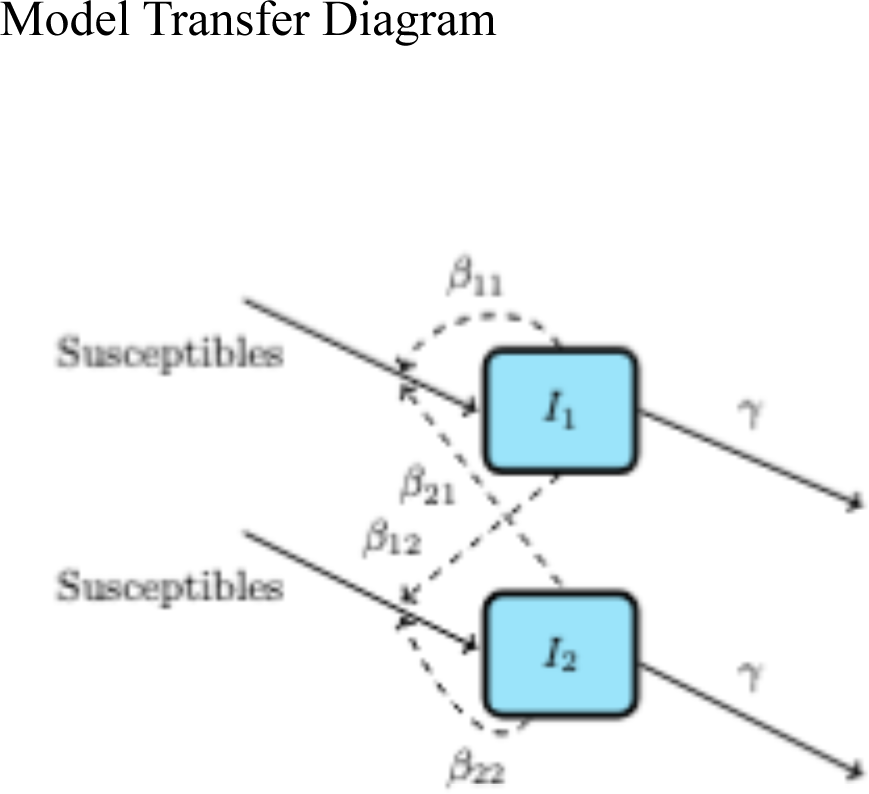
Model transfer diagram for two-state infectious interactions, exponential growth phase only. I1 = Adults; I2 = Children. Susceptible individuals are considered unlimited; γ represents the rate of becoming non-infectious.

**Figure S3.**
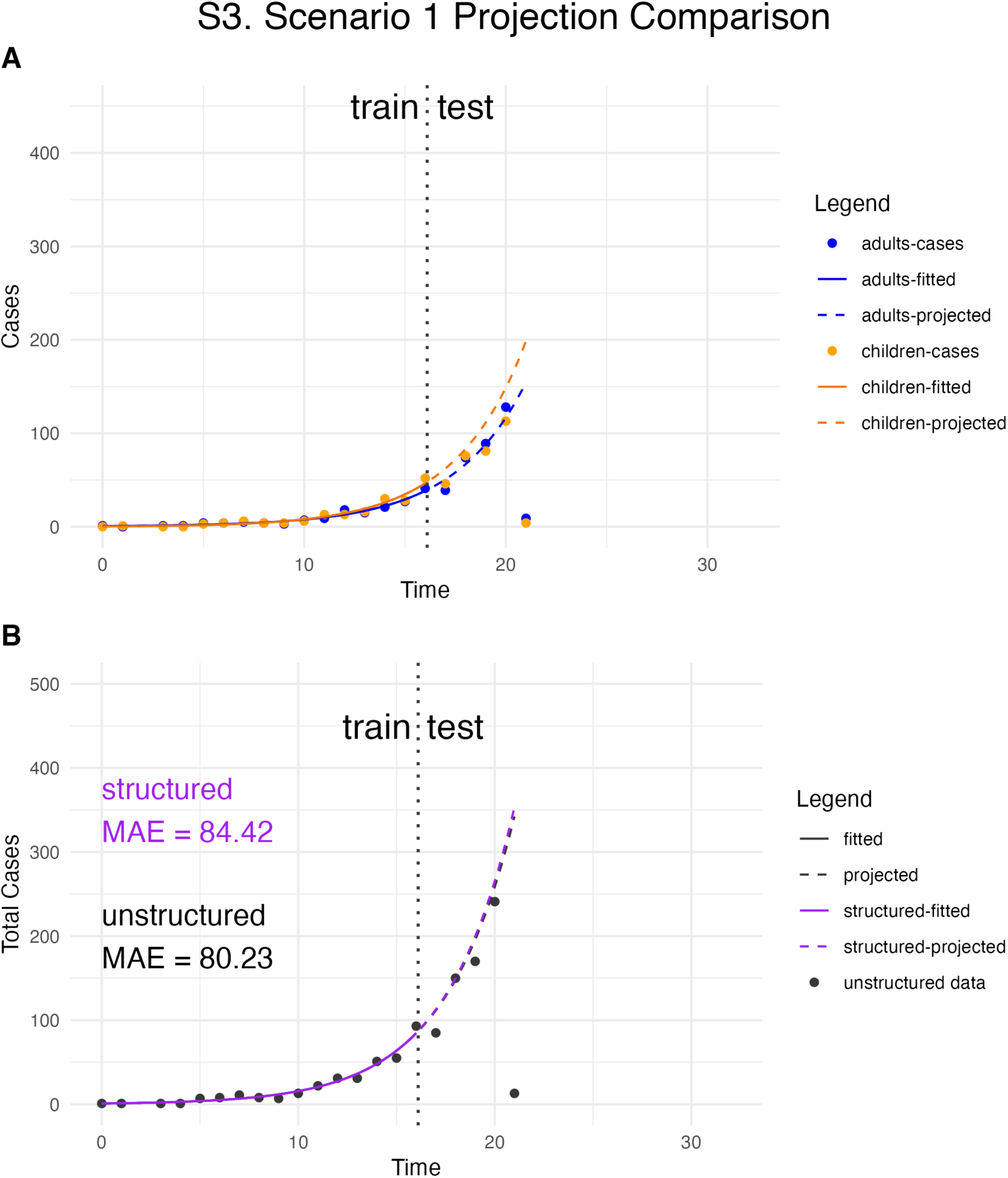
Case time series projection: *Scenario 1 random sample* out of 200 replicates. Left of the dotted vertical line, the models are fit to the training data sets. Right of the dotted line, the models predict the trends of infection spread in the testing (holdout) data. Panel A shows the structured data and model; Panel B shows the unstructured data (black dots) and model (blue line). In Panel B, the purple solid line represents the sum of the two age groups at each time point from the structured model fit; the purple dashed line represents the projected sum of the two age groups from the structured model. MAE stands for Mean Absolute Error.

**Figure S4.**
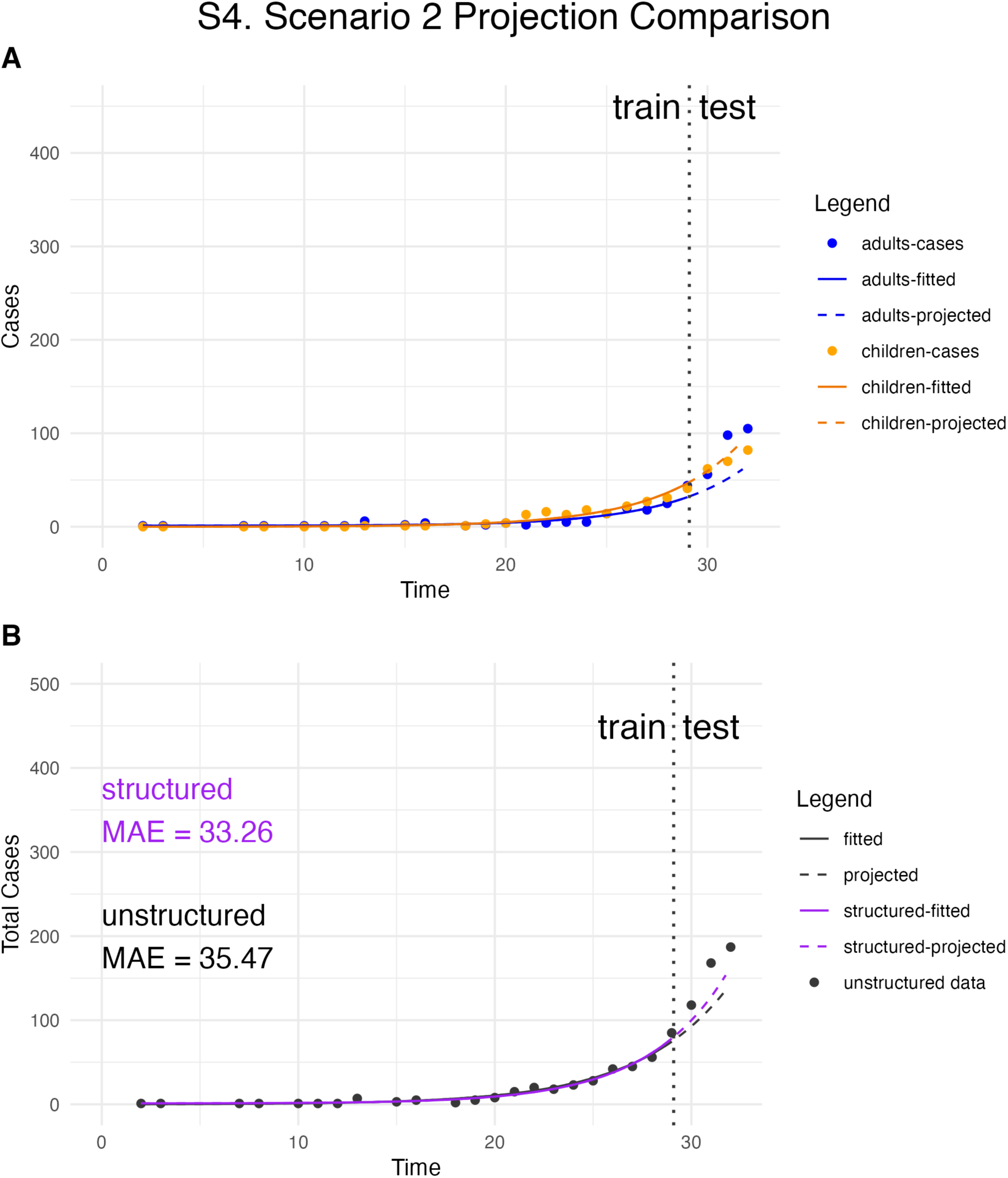
Case time series projection: *Scenario 2 random sample out of 200 replicates.* Left of the dotted vertical line, the models are fit to the training data sets. Right of the dotted line, the models predict thetrends of infection spread in the testing (holdout) data. Panel A shows the structured data and model; Panel B shows the unstructured data (black dots) and model (blue line). In Panel B, the purple solid line represents the sum of the two age groups at each time point from the structured model fit; the purple dashed line represents the projected sum of the two age groups from the structured model. MAE stands for Mean Absolute Error.

**Figure S5.**
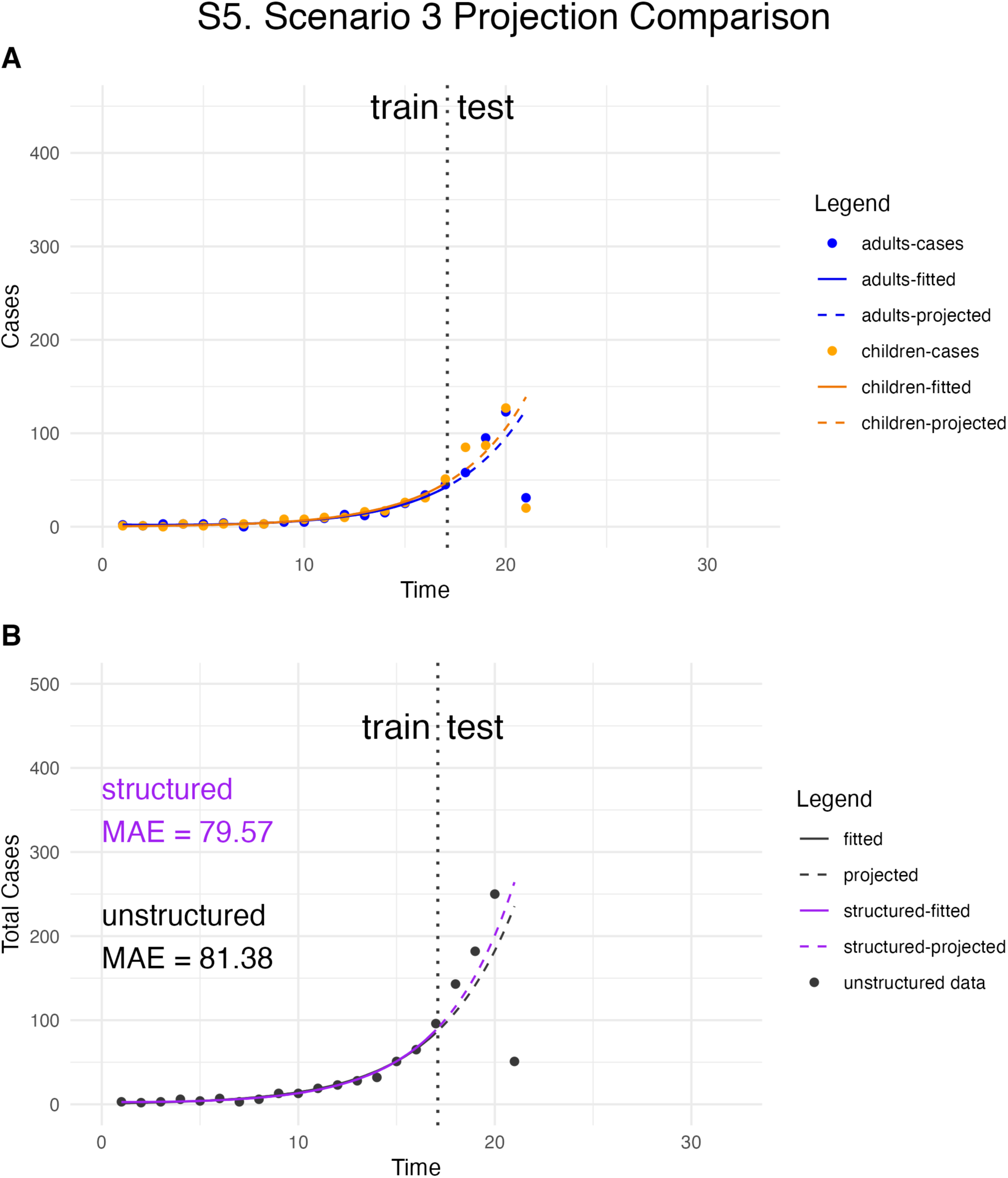
Case time series projection: *Scenario 3 random sample out of 200 replicates*. Left of the dotted vertical line, the models are fit to the training data sets. Right of the dotted line, the models predict the trends of infection spread in the testing (holdout) data. Panel A shows the structured data and model; Panel B shows the unstructured data (black dots) and model (blue line). In Panel B, the purple solid line represents the sum of the two age groups at each time point from the structured model fit; the purple dashed line represents the projected sum of the two age groups from the structured model. MAE stands for Mean Absolute Error.

**Figure S6.**
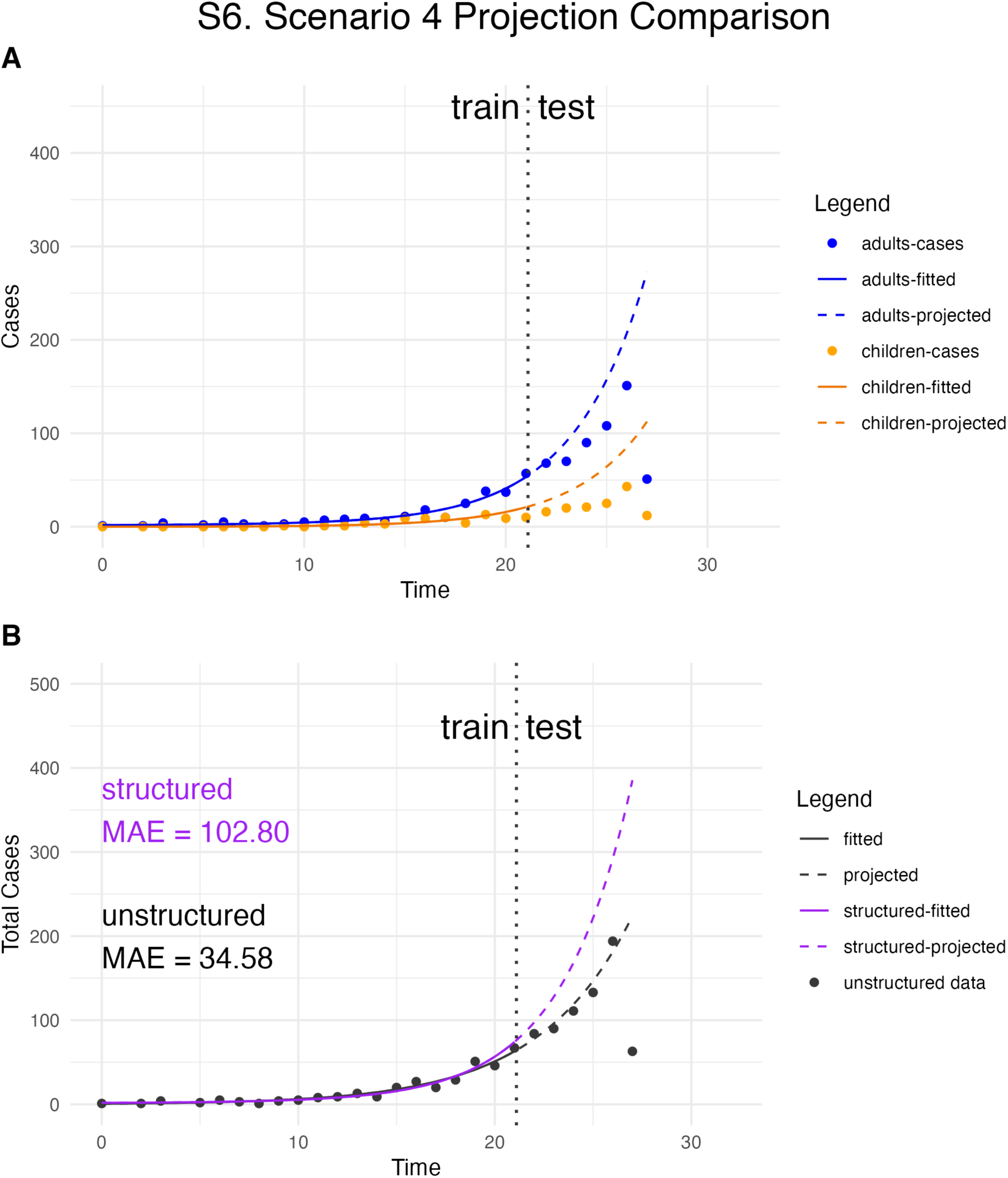
Case time series projection: *Scenario 4 random sample out of 200 replicates.* Left of the dotted vertical line, the models are fit to the training data sets. Right of the dotted line, the models predict the trends of infection spread in the testing (holdout) data. Panel A shows the structured data and model; Panel B shows the unstructured data (black dots) and model (blue line). In Panel B, the purple solid line represents the sum of the two age groups at each time point from the structured model fit; the purple dashed line represents the projected sum of the two age groups from the structured model. MAE stands for Mean Absolute Error.

